# Source-linked multispecialty clinical reasoning and tumour-board decision-making in colorectal cancer

**DOI:** 10.64898/2026.10.19.26365647

**Authors:** Victor A. Gombolevskiy, Marina I. Sekacheva, Varvara D. Sanikovich, Nikolay N. Bagmet, Daiana A. Balaeva, Nikolay B. Nechaev, Aleksandr I. Nesterov, Alexey A. Bokov, Ivan I. Panin, Iaroslav R. Bespalov, Anastasia V. Gribakina, Dmitrii V. Dylov, Ivan V. Oseledets

## Abstract

Clinical records preserve patient findings and final recommendations but rarely capture how specialists select evidence, develop alternative interpretations, disagree, and converge on a management strategy. We describe a dataset derived from ten retrospectively selected, completed cases of histologically confirmed colorectal cancer. For each case, an administrator uploaded de-identified textual clinical records to an authenticated web-based annotation platform. A medical oncologist, surgical oncologist, and radiation oncologist, each with more than ten years of clinical experience, independently linked relevant source fragments to patient-specific interpretations and documented clinical problems, differential hypotheses, uncertainty, and diagnostic and treatment strategies. After the three records were locked, a large language model generated candidate questions for review by the tumour-board chair. The four physicians then discussed each case online; recordings were automatically transcribed and verified by two medical residents. The dataset comprises 30 source-linked specialist reasoning records, 10 verified case-level discussion transcripts, 10 final tumour-board decisions, and associated provenance and quality-control metadata. It supports research on evidence-grounded reasoning, cross-specialty disagreement, multidisciplinary decision-making, and multi-agent clinical decision support.

## 1 Background & Summary

Contemporary medical artificial intelligence systems are commonly developed using medical images, structured clinical variables, laboratory results, clinical notes, diagnoses, treatment records and outcomes. These data preserve observable inputs and endpoints but often omit how clinicians select relevant evidence, assign patient-specific significance to individual findings, consider alternative interpretations, calibrate uncertainty and connect hypotheses to proposed actions. Models trained predominantly on conventional medical records may therefore learn associations between available patient data and recorded decisions without access to the externalised reasoning that linked them.

Clinical reasoning cannot be equated with written text or treated as a complete record of a clinician’s internal cognitive process. Some decisions arise rapidly through pattern recognition and accumulated experience, whereas others involve deliberate comparison of competing hypotheses, contradictory evidence and potential benefits and harms. Neither process can be observed in full. The present dataset therefore records externalised clinical reasoning: facts, interpretations, hypotheses, counterarguments, uncertainties and conditional decisions that physicians explicitly documented or articulated during multidisciplinary discussion.

To frame which components of clinical reasoning can be represented as reusable data, we used a conceptual framework based on the observability of reasoning. Clinical reasoning was considered as a continuum from largely unobservable internal processes, including rapid pattern recognition, deliberate analytical reasoning, and metacognitive error checking, through spoken externalisation in clinical dialogue and multidisciplinary discussion, to increasingly persistent written representations ranging from conventional clinical documentation to expanded and structured reasoning records. Previous work on clinical reasoning supports the distinction between rapid non-analytical and deliberate analytical processes, although their interaction is dynamic and context dependent [1, 2].

The framework is not intended as a validated cognitive taxonomy. Rather, it was used to guide data capture by distinguishing internal reasoning, which cannot be directly observed, from progressively more explicit external representations. Dataset version 1.0 samples several observable levels of this continuum by linking conventional source clinical records, structured specialist reasoning, recorded multidisciplinary discussion, and a final tumour-board decision (Figure 1).

**Figure 1:**
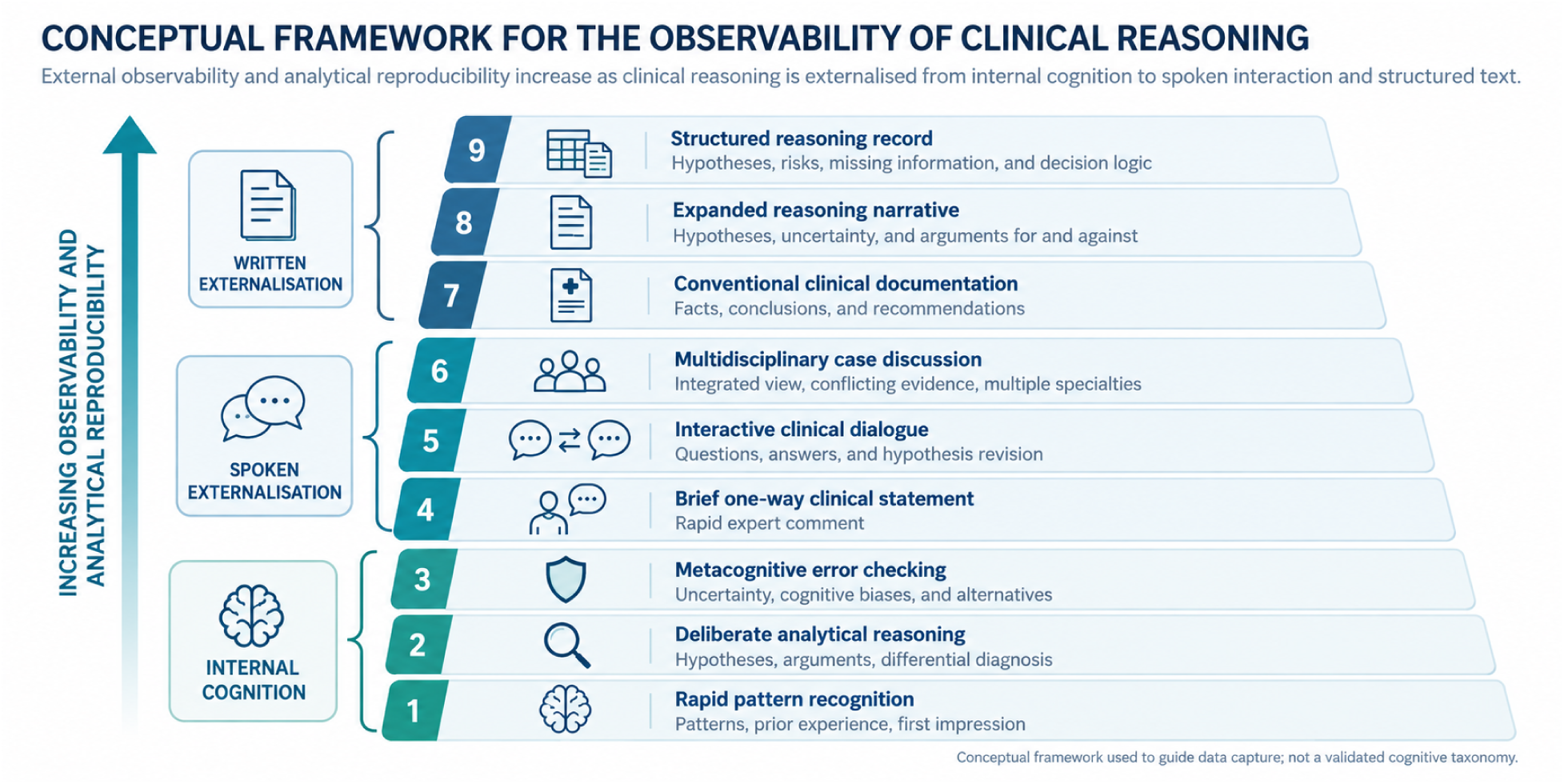
Conceptual framework for the observability of clinical reasoning. Clinical reasoning ranges from largely unobservable internal cognitive processes to spoken and written external representations that can increasingly be preserved, reproduced, and analysed. The nine-level framework distinguishes internal cognition, spoken externalisation, and written externalisation. Dataset version 1.0 captures several observable levels of this continuum through source clinical documentation, structured specialist reasoning, recorded multidisciplinary discussion, and final tumour-board decisions. The framework describes progressively more explicit external representations and is not intended as a validated cognitive taxonomy.

Multidisciplinary tumour boards are intended to integrate complementary professional expertise. In colorectal cancer, the completeness and quality of clinical information influence whether a board can reach a decision, and multidisciplinary review may modify an initially proposed management plan. The contribution of individual participants, clinical hierarchy and the organisation of discussion can also influence decision-making [3–5]. Conventional tumour-board documentation, however, usually preserves the final recommendation more completely than the independent positions, disputed assumptions and arguments through which that recommendation was reached.

Several recent medical datasets have moved beyond simple input–label pairs by preserving intermediate expert information and explicit links between derived interpretations and supporting evidence. Examples include chest radiographs paired with dictation audio and gaze trajectories, dialogue-to-note corpora, medical question-answer datasets with evidence attribution, electronic-health-record datasets with sentence-level relevance labels, and expert-verified multilevel reasoning annotations for breast ultrasound [6–11]. Recent work has also described clinician-reviewed large-language-model assistance during medical annotation [12], while CheXthought demonstrates the feasibility of collecting multiple human reasoning traces around the same chest radiograph [13]. These resources primarily represent reasoning around an imaging examination, a clinical encounter or a question-answer pair. The present dataset shifts the unit of analysis to a completed patient-level oncological case reviewed independently by several treatment specialties and subsequently discussed jointly.

The closest resources are three reasoning-oriented benchmarks for language models. DiReCT contains 511 clinical notes in which physicians annotated the path of diagnostic reasoning from individual observations in the note to the final diagnosis, supported by a diagnostic knowledge graph [14]; its annotation is restricted to the diagnostic task and does not capture the comparison of positions between specialists of different profiles. MedCaseReasoning comprises 14,489 diagnostic question–answer pairs built from open-access published case reports, each paired with statements of clinicians’ diagnostic reasoning derived from the publication [15]; the reasoning is thus reconstructed retrospectively from the authors’ narrative rather than recorded independently by several specialists for the same case. MTBBench simulates molecular tumour board decision-making on multimodal, longitudinal oncology data and uses clinician-validated reference answers to evaluate agentic systems based on large language models [16]; it is conceived as a benchmark with reference answers rather than as a record of the discussion process itself. In contrast, the present dataset records the independent positions of three specialists of different profiles on the same case linked to source documents, the explicit disagreements between them, the course of the spoken tumour-board discussion and the final decision formulated by the chair.

The dataset connects patient-level source documents, selected source spans, three independently generated specialty-specific reasoning records, candidate questions derived from differences between those records, a verified case-level multidisciplinary discussion, and a final tumour-board decision formulated by the chair. Independent records were completed and locked before specialists could inspect one another’s annotations. The resulting reusable object is therefore not only a clinical record or final recommendation, but a traceable sequence from source evidence to individual interpretation, professional disagreement, multidisciplinary discussion, and a chair-formulated final decision.

This representation enables investigation of source-grounded clinical reasoning, cross-specialty variability, unsupported claims, uncertainty, changes in professional position after discussion and multi-agent decision-support methods that maintain distinct perspectives. The dataset does not establish that the final tumour-board decision was clinically superior to the original treatment decision, does not measure patient outcomes and is not intended to serve as a standalone training corpus for a general-purpose clinical model. Patient preferences and patient-defined benefit–harm trade-offs were not captured as a separate structured layer in this release.

**Figure 2:**
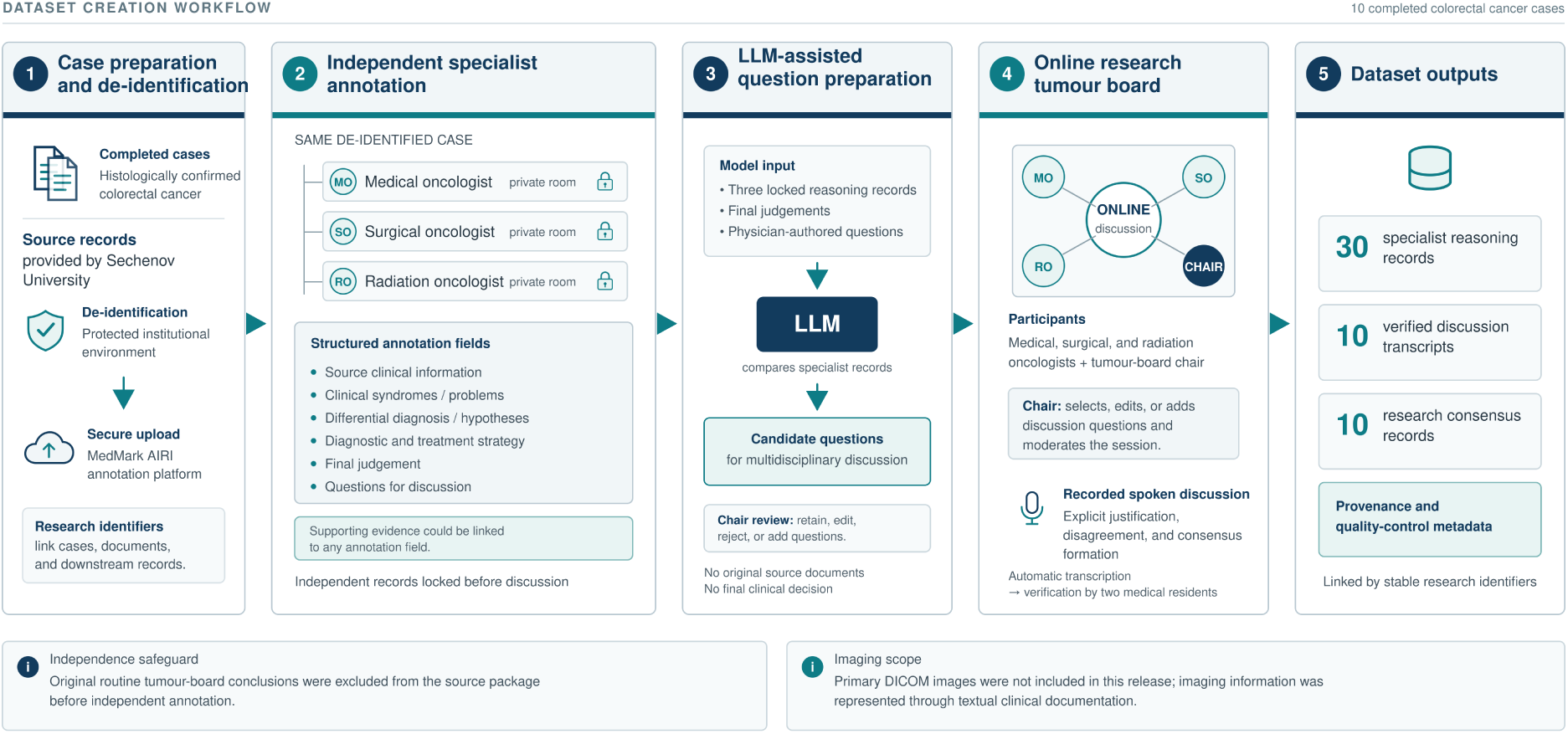
Dataset creation workflow. (1) Completed colorectal cancer cases and source clinical records provided by Sechenov University were de-identified within the protected institutional environment and uploaded to the MedMark AIRI annotation platform. (2) A medical oncologist, surgical oncologist, and radiation oncologist independently annotated the same de-identified case in separate private annotation rooms; the completed records were locked before multidisciplinary discussion. (3) A large language model compared the three locked specialist reasoning records and generated candidate questions for multidisciplinary discussion. The model had no access to the original source documents and did not make the final clinical decision. Candidate questions were reviewed by the tumour-board chair. (4) The three specialists and the chair participated in recorded online research tumour boards. The recordings were automatically transcribed and subsequently verified by two medical residents. (5) The resulting dataset comprised 30 specialist reasoning records, 10 verified multidisciplinary discussion transcripts, 10 final tumour-board decisions, and associated provenance and quality-control metadata. Original routine tumour-board conclusions were excluded from the source packages before independent annotation. Primary DICOM images were not included in this release; imaging information was represented through textual clinical documentation.

## 2 Methods

### 2.1 Study design

This study was a retrospective data-curation and expert-annotation project based on previously completed clinical cases. All source clinical records were provided by Sechenov University, Moscow, Russian Federation. The original diagnostic work-up, treatment, and routine multidisciplinary tumour-board review had been completed before the research annotation process.

To preserve the independence of the research annotations, records and conclusions from the original routine tumour-board discussions were excluded from the source packages provided to the annotators. The specialists therefore reviewed the remaining clinical documentation without access to the previous multidisciplinary tumour-board decision.

The repeated expert assessment described in this Data Descriptor was performed exclusively for dataset construction. It did not constitute prospective clinical care, did not alter patient management, and had no influence on the treatment or outcomes of the included patients.

Ten completed cases of histologically confirmed colorectal cancer were included. Expert annotation and research tumour-board discussions were conducted between February and August 2026 by physicians affiliated with three medical centres: Sechenov University, Moscow, Russian Federation; Petrovsky National Research Center of Surgery, Moscow, Russian Federation; and Central Clinical Hospital “RZD-Medicine”, Moscow, Russian Federation.

The version described in this manuscript represents a fixed ten-case dataset scope. Subsequent cases, extensions of the annotation framework, and downstream use of the dataset for the development or evaluation of artificial intelligence systems are outside the scope of the present Data Descriptor.

### 2.2 Selection of clinical cases

Cases were purposively selected by clinicians from Sechenov University; the tumour-board chair did not participate in case selection. Selection was based on the suitability of each case for substantive multidisciplinary review rather than on a formal measure of case complexity.

Eligible cases were required to contain a diagnostic or therapeutic question for which assessment by medical oncology, surgical oncology, and radiation oncology could provide complementary professional perspectives. Cases were additionally required to have sufficiently complete digital clinical documentation to reconstruct the relevant diagnostic and treatment trajectory.

Inclusion criteria were:

- histologically confirmed colorectal cancer;
- completion of the relevant clinical episode;
- sufficient digital clinical documentation to reconstruct the diagnostic and treatment trajectory;
- presence of a diagnostic or therapeutic question suitable for substantive assessment by the three participating specialties;
- previous completion of a routine multidisciplinary tumour-board review at Sechenov University;
- documented patient permission for secondary use of the clinical data for scientific research, including controlled sharing of de-identified data; and
- feasibility of de-identification before transfer to the research annotation platform.

Exclusion criteria were:

- absence of key clinical documentation;
- inability to reconstruct the sequence of clinically relevant events;
- absence of a substantive diagnostic or therapeutic question requiring multidisciplinary assessment;
- duplication of a previously included clinical episode; or
- inability to prepare the clinical material in a de-identified form suitable for the intended research workflow.

The sample was purposive rather than consecutive and was not designed to estimate the prevalence of clinical scenarios, treatment strategies, or outcomes among patients with colorectal cancer. Although every included case had previously undergone routine multidisciplinary review at Sechenov University, the records and conclusions of those original tumour boards were deliberately excluded from the source packages provided to the research annotators.

### 2.3 Source clinical records

All source clinical records were provided by Sechenov University. For each included case, the source package was assembled from the available digital medical documentation generated during routine clinical care.

Depending on the individual clinical trajectory, the package could include discharge summaries, histories of the present illness, laboratory results, endoscopy reports, computed tomography and magnetic resonance imaging reports, pathology reports, immunohistochemistry results, molecular genetic test results, records of surgical treatment, systemic anticancer therapy and radiation therapy, response-assessment reports, and follow-up documentation.

The source package was intended to preserve the clinical information that had been available during the completed episode of care and that could contribute to retrospective multidisciplinary assessment. Records and conclusions from the original routine tumour-board discussion were deliberately excluded before the package was provided to the research annotators in order to prevent the previous multidisciplinary decision from influencing their independent assessments.

Each case was assigned a unique research identifier, and each source document was assigned a document-level identifier linked to the corresponding case. These identifiers were used throughout the annotation workflow to connect source documents, selected source fragments, specialist interpretations, multidisciplinary discussion, and the resulting final tumour-board decision.

Original Digital Imaging and Communications in Medicine (DICOM) examinations were not included in this dataset. Imaging information was represented through the corresponding textual radiology reports and other textual clinical documentation. Extension of the data model to primary medical images is outside the scope of the present release.

### 2.4 De-identification and disclosure-risk management

De-identification was performed by authorised personnel at Sechenov University within the institution’s protected information environment before any clinical material was transferred to the research annotation platform. Identifiable source records were not provided directly to the external annotators or research team.

Direct personal identifiers were removed before upload, including patient names, dates of birth, postal addresses, telephone numbers and email addresses, medical-record numbers, health-insurance identifiers, national insurance identifiers, and other document or account numbers that could directly identify an individual. Names of healthcare organisations appearing in the clinical documentation were replaced with neutral labels where required for de-identification.

Original dates of clinical events were retained. Temporal relationships between diagnostic procedures, treatment, response assessment, disease progression, and follow-up constituted clinically relevant information for retrospective reasoning and were therefore preserved rather than converted to relative intervals.

After the initial de-identification procedure, the prepared case package underwent an additional review by an independent oncologist before annotation; in addition, the completeness of identifier removal was repeatedly re-checked by several physicians. Administrative access to the MedMark AIRI platform was granted to an authorised Sechenov University staff member, who uploaded the de-identified source documents directly to the platform, assigned the cases to the predefined annotation workflow, and verified that the uploaded materials corresponded to the intended research case.

Owing to this repeated checking, the risk of re-identification was minimised and, for a dataset of ten cases, is considered to be very low. Before public dataset release, the complete de-identified dataset will undergo an additional disclosure-risk audit covering free-text content, document metadata, indirect identifiers, internal technical identifiers, and combinations of clinical characteristics that could increase the risk of re-identification. Only the version approved for scientific data sharing under controlled access will be deposited as the version of record.

### 2.5 Selection of clinical experts

Each case was independently reviewed by the same three physicians representing complementary treatment specialties: one medical oncologist, one surgical oncologist, and one radiation oncologist.

Eligibility for the three annotating specialists required more than ten years of active clinical practice in the relevant specialty. The experts were affiliated with three different medical centres: Sechenov University, Petrovsky National Research Center of Surgery, and Central Clinical Hospital “RZD-Medicine”. The multidisciplinary composition was selected to ensure that the same patient-level clinical information was assessed from distinct professional perspectives relevant to systemic therapy, surgical management, and radiation treatment.

The same three specialists independently reviewed all ten cases. Each physician received an individual authenticated account and a separate specialty-specific annotation room within the MedMark AIRI platform. During the independent annotation stage, specialists did not have access to the annotations, conclusions, or discussion questions generated by the other experts. Their records were locked after completion and became visible to the other participants only when the research tumour-board stage began.

A fourth physician served as the chair of the research tumour board. The chair did not participate in case selection or in the independent annotation stage and joined the workflow only after the three specialist records had been completed. During the tumour-board sessions, the chair reviewed the prepared discussion questions, moderated the discussion, invited the specialists to justify or reconsider their positions, and subsequently formulated the final tumour-board decision for each case on the basis of the multidisciplinary discussion.

Annotator names and contact details will not be included in the Dataset version 1.0 release. The co-authors and annotators did not take part in the treatment of the patients whose cases were included in the dataset.

### 2.6 Annotation platform and administrative workflow

Annotation was performed using the custom authenticated MedMark AIRI web-based annotation platform (version 1.0). The platform implemented role-based access for an administrator, specialist annotators, and the research tumour-board chair. MedMark AIRI functioned as an annotation, provenance-linking, and workflow-management instrument and did not generate or modify the physicians’ clinical interpretations.

The administrator role was assigned to an authorised employee of Sechenov University. Through a dedicated administrative panel, the administrator uploaded the previously de-identified clinical materials, created individual authenticated user accounts, assigned professional roles, and allocated case-specific annotation tasks to each physician according to the study protocol.

The platform allowed case assignments to be configured independently for each user. In the present study, the same ten cases were assigned separately to each of the three specialist annotators, resulting in 30 independent specialist–case annotation tasks. Physicians could complete their assigned cases asynchronously and independently at any time during the individual annotation stage.

Each specialist worked within a private role-specific annotation environment and had no access to the other physicians’ annotations while preparing the case independently. The interface combined a viewer for the source clinical documents with a structured annotation panel. The annotation sections acted as cognitive scaffolding, prompting physicians to identify clinically relevant source information, interpret its patient-specific significance, formulate clinical problems and alternative hypotheses, and describe diagnostic and treatment strategies. The structured fields defined the organisation of the resulting data but did not prescribe the clinical conclusion or require agreement between specialists.

Within the source-document viewer, physicians could select clinically relevant source fragments and associate them with their own written interpretations. This design allowed the resulting annotation to preserve an explicit link between source evidence and the corresponding physician reasoning.

After all three specialists had completed and locked their independent annotations for the same case, the case became eligible for multidisciplinary discussion. The administrator coordinated the availability of the three specialists and the tumour-board chair and scheduled the corresponding online research tumour-board session. The detailed tumour-board and recording procedures are described below.

### 2.7 Unit of annotation

The annotation framework was organised at two linked levels: an atomic evidence-linked annotation unit and a case-level specialist reasoning record.

The atomic annotation unit consisted of a clinically relevant evidence fragment linked to a physician-generated interpretation. Evidence could originate from the patient-level source clinical records or, where clinically relevant, from external supporting materials used by the physician, such as clinical guidelines or other reference sources. Annotators were able to attach supporting evidence to any structured annotation field, including higher-level reasoning sections, diagnostic conclusions, recommendations, and the final individual judgement.

For patient-derived evidence, annotators were instructed to select the minimum source fragment required to support the corresponding clinical statement rather than reproduce entire documents. The linked physician comment was intended to explain the patient-specific significance of the selected evidence and, when applicable, how that evidence supported or weakened a clinical hypothesis, affected a diagnostic or treatment decision, or justified the absence of an additional action.

The general reasoning sequence underlying the annotation framework was:

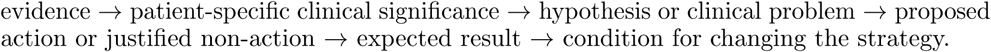

This sequence was used as a structuring principle rather than as a requirement that every annotation contain all six elements. The depth of an individual annotation depended on the clinical relevance, uncertainty, and decision-making consequences of the underlying evidence.

Evidence and physician interpretation were stored as distinct but explicitly linked components. This distinction preserved provenance and allowed subsequent users to differentiate information present in the original clinical record, external evidence consulted by the physician, and the specialist’s own interpretation or inference. A clinical statement could therefore be traced back to the evidence used to support it.

At the second level, all annotations produced by one specialist for one patient case were combined into a complete specialist reasoning record. This record contained source-linked interpretations, synthesis of clinical problems, differential hypotheses, diagnostic and treatment strategy, individual diagnosis, recommendations, final judgement, and questions for multidisciplinary discussion. The dataset therefore contains 30 case-level specialist reasoning records generated from ten patient cases reviewed independently by the same three specialists.

**Figure 3:**
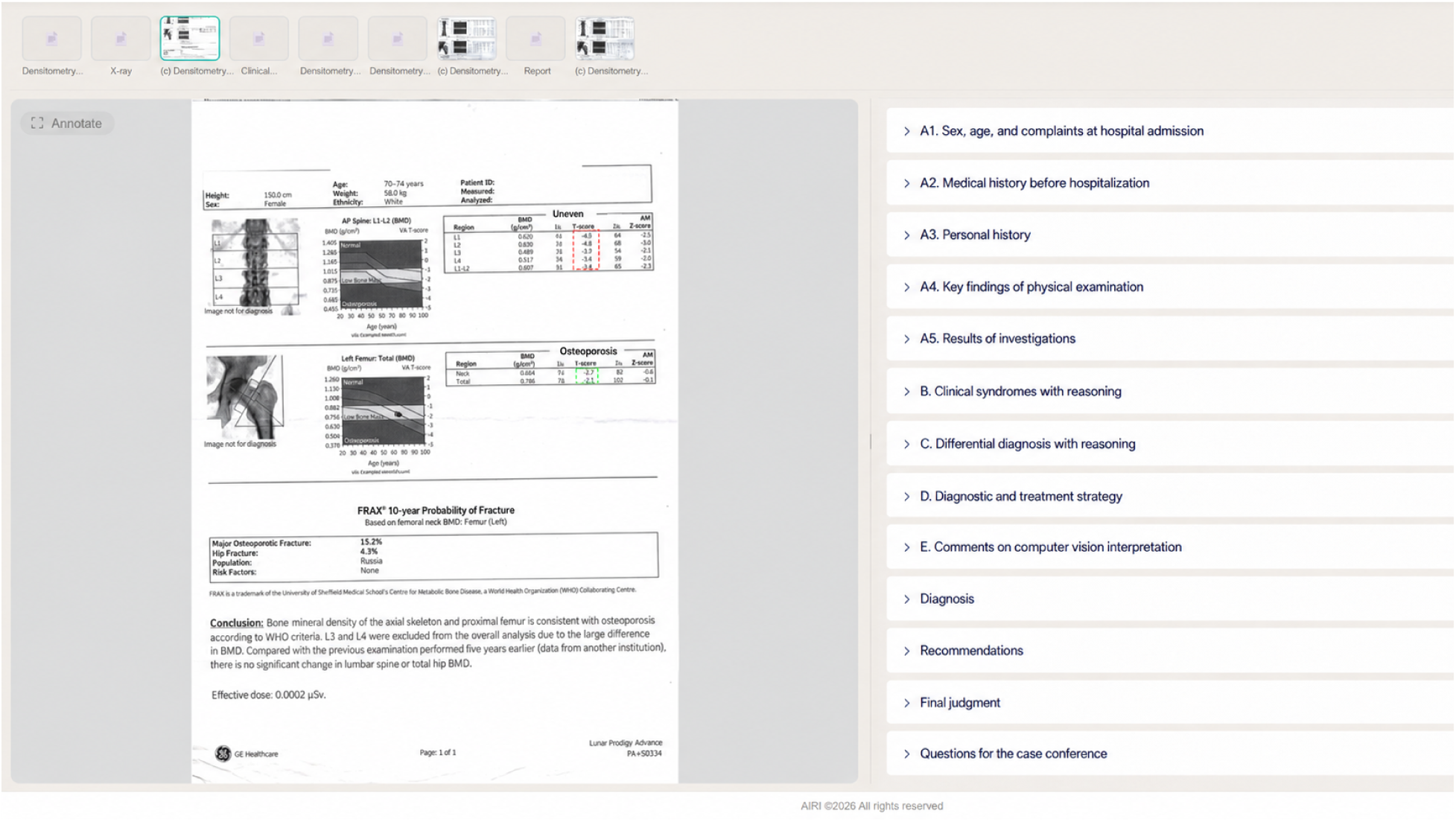
Specialist-facing MedMark AIRI annotation interface illustrated using a non-study densitometry example. Source files preloaded by the administrator are displayed in the upper file ribbon. The left panel shows the active source document; physician-selected evidence fragments are highlighted in colour and linked to the corresponding annotations. The right panel contains the structured annotation workflow, which the specialist completes sequentially from fields A1–A5 through sections B–D and the concluding fields Diagnosis, Recommendations, Final judgement, and Questions for multidisciplinary discussion. The screenshot illustrates the functionality of the annotation platform and does not represent a patient included in Dataset version 1.0. The optional computer-vision field visible in the generic interface was not used in the present study.

### 2.8 Structured annotation fields

The annotation interface organised each specialist’s reasoning into a predefined sequence of structured fields. The purpose of this structure was to encourage explicit documentation of the transition from source clinical information to interpretation, competing hypotheses, and patient-specific management decisions, while preserving the specialist’s freedom to reach an independent clinical conclusion. Supporting evidence could be linked to any of the annotation fields.

Section A: source clinical information and patient-specific interpretation. Section A contained five fields designed to identify clinically relevant information from the source records and document its significance for the individual case.

- A1. Sex, age, and presenting complaints. The specialist identified relevant demographic and symptomatic information and documented its significance for the current clinical problem.
- A2. History of the present illness. The specialist reconstructed the clinically relevant temporal sequence of diagnosis, treatment, response, progression, and other events contributing to the current decision.
- A3. Relevant medical history. The specialist selected comorbidities, previous interventions, treatment exposures, and other background factors that could modify diagnosis, treatment selection, expected benefit, or risk.
- A4. Key objective findings. The specialist documented clinically relevant physical, functional, or other objective findings where these were available in the source material and contributed to the decision.
- A5. Results of investigations. The specialist selected and interpreted relevant laboratory, endoscopic, radiological, pathological, immunohistochemical, molecular, and other diagnostic results.

Section A was not intended to reproduce the complete medical record. Physicians selected information that they considered relevant to the clinical problem and could associate each selected source fragment with a patient-specific interpretation explaining why the information mattered for subsequent reasoning.

Section B: clinical syndromes and problem representation. Section B required the specialist to integrate individual findings into higher-level clinical syndromes or patient-specific clinical problems. The physician could link these interpretations to supporting patient records or external evidence and describe the findings supporting the problem representation, the proposed clinical mechanism, relevant uncertainty, and alternative interpretations when applicable.

Section C: differential diagnosis and competing hypotheses. Section C captured the set of clinically plausible hypotheses considered by the specialist. Physicians could document the relative probability of each hypothesis, evidence supporting or arguing against it, information required to confirm or reject it, and clinically important alternatives that should not be overlooked despite lower estimated probability.

In patients with an already histologically confirmed malignancy, differential reasoning was not limited to the identity of the primary tumour. It could also concern the interpretation of new lesions, causes of complications, patterns of progression or treatment response, resectability, and other uncertainties capable of changing the subsequent management strategy.

Section D: diagnostic and treatment strategy. Section D captured the specialist’s proposed patient-specific management pathway. Physicians were encouraged to document the clinical objective, proposed diagnostic or therapeutic action, order and timing of interventions, expected result, conditional branches of the strategy, monitoring of effectiveness and safety, and the circumstances under which the proposed plan should be reconsidered.

The field also allowed specialists to document a justified decision not to perform an additional investigation or intervention when the available evidence did not reach the threshold for further action.

Concluding fields. After completing the structured reasoning sections, each specialist recorded an individual diagnosis or problem formulation, recommendations, a concise final judgement, and questions for multidisciplinary discussion.

The final judgement provided a concise representation of the specialist’s position after reviewing the complete case, whereas the questions for multidisciplinary discussion identified unresolved diagnostic or therapeutic dilemmas for which input from the other specialties could potentially modify the proposed strategy.

Because supporting evidence could be attached throughout the interface, the resulting specialist record combined lower-level source-linked interpretations with higher-level synthesis, hypotheses, management decisions, and an explicit summary of unresolved questions.

All structured annotation fields used in the present study were mandatory. Fields were not left blank; when information was unavailable, not applicable, or could not be assessed within the specialist’s role, the annotator was required to explicitly document the reason.

The general MedMark AIRI platform also contained an optional field for assessment of computer-vision outputs. This functionality was not used in the present dataset because the ten colorectal cancer cases were analysed exclusively from textual clinical materials and did not include primary medical images or computer-vision results. Computer-vision-assisted annotation is outside the scope of the present release.

### 2.9 Annotation rounds

Data collection was organised in three sequential annotation rounds. The first round comprised cases 001–003, the second comprised cases 004–006, and the third comprised cases 007–010.

During each round, the same three specialists independently reviewed the same set of patient cases. Annotation was performed asynchronously within the physicians’ private annotation rooms, typically over the course of a working week. Completion of one case required up to approximately two hours per specialist.

Once all three independent specialist records for the cases included in a round had been completed and locked, the corresponding cases became eligible for multidisciplinary discussion. The administrator then coordinated the availability of the three specialists and the tumour-board chair and scheduled an online research tumour-board session.

After each completed round, the annotators received general, non-personalised feedback from the AIRI project lead regarding the explicitness and structural completeness of the documented clinical reasoning. The feedback focused on whether hypotheses, supporting and opposing evidence, uncertainty, conditional decision logic, and monitoring criteria were expressed clearly enough to be observable in the resulting text. It did not assess physician competence, rank specialists, or provide case-specific clinical answers.

Because feedback was provided between rounds, the annotation process was intentionally iterative. Later records may therefore reflect increasing familiarity with the annotation framework and greater explicitness of written reasoning. This feature of the data-generation process should be considered when comparing annotations across sequential cases.

Across the three rounds, the procedure generated 30 independent specialist–case reasoning records from ten patient cases before multidisciplinary discussion.

### 2.10 Large-language-model-assisted preparation of discussion questions

After completion and locking of the three independent specialist annotations for a case, a large language model (LLM) was used to compare the three reasoning records and generate candidate questions for multidisciplinary discussion.

The model received the complete annotation records produced by the medical oncologist, surgical oncologist, and radiation oncologist for the same patient case, including their reasoning sections, individual final judgements, and physician-authored questions for the tumour board. The model did not receive the original patient source documents, clinical guidelines, or other external clinical reference materials at this stage.

Across the data-collection period, the LLM-assisted step was performed using ChatGPT Codex 5.4/5.5 extra-high (OpenAI) through the standard web interface, with the model configuration available at the time of each round. All patient-level material provided to the model had previously undergone the de-identification procedure described above. Privacy settings were applied before data were submitted; no personal patient data were transmitted to ChatGPT (OpenAI), and processing in this mode was considered permissible for research purposes. The exact prompt used to generate candidate discussion questions is preserved and will be provided with the accompanying methodological materials.

The purpose of the LLM was not to determine the correct diagnosis or treatment strategy, but to identify clinically meaningful differences between the independently documented professional positions. The prompt encouraged identification of conflicting interpretations, differences in proposed diagnostic or treatment strategies, insufficiently supported assumptions, unresolved uncertainty, missing arguments, and benefit–harm trade-offs that could materially affect the multidisciplinary decision.

The LLM was specifically used to formulate questions capable of eliciting explicit justification of competing positions during the subsequent discussion. This design was intended to reduce the likelihood that disagreement would remain implicit or that an initial position would be accepted without examination solely because of professional seniority or conversational dynamics. The model was not provided with information about the relative status or seniority of the three specialists when generating these questions.

Before each research tumour-board session, the administrator sent the generated candidate questions to the tumour-board chair. The chair retained full editorial and clinical control over their use and could retain, reformulate, omit, or replace any model-generated question and could introduce additional questions independently. The final selection and wording of questions used during the meeting were therefore determined by the chair.

The LLM did not participate in the independent annotation stage, did not modify the physicians’ locked records, did not participate as an independent clinical agent during the tumour board, and did not generate the final tumour-board decision. Clinical responsibility for the multidisciplinary discussion remained with the participating physicians, while the tumour-board chair was responsible for formulating the final decision.

The original prompt used for this LLM-assisted step has been retained. However, the complete set of intermediate model-generated candidate questions and the chair’s individual retain–modify–reject decisions were not systematically archived during data collection. These intermediate outputs are therefore not included as a separately reusable data layer in the present release.

### 2.11 Online research tumour boards

Three online research tumour-board sessions were conducted after completion of the independent annotation rounds. The first and second sessions each considered three patient cases, and the third session considered four cases. Each session lasted approximately 1.5–2 hours.

Each research tumour board included the same three specialist annotators—the medical oncologist, surgical oncologist, and radiation oncologist—together with the tumour-board chair. The chair joined the workflow only at the multidisciplinary discussion stage and did not participate in the preceding independent annotations.

Before the meeting, all three specialist records for the corresponding cases had been completed and locked. During the tumour-board stage, participants were able to review the other specialists’ documented positions. The chair additionally received the candidate discussion questions prepared through the LLM-assisted procedure described above and retained full discretion to select, reformulate, omit, or supplement these questions.

Meetings were conducted using SaluteJazz (version 26.61.10; Sber), a videoconferencing service supporting meeting recording and speaker-attributed automatic speech transcription.

The multidisciplinary stage was intentionally conducted as a spoken discussion rather than as continued written annotation. This format was selected to minimise interruption of clinical reasoning by manual text entry and to preserve the temporal sequence of arguments, counterarguments, clarifications, and changes in professional position. Participants could justify their initial recommendations, challenge assumptions made by other specialists, introduce additional evidence, and revise their own positions during discussion.

The online sessions were recorded for subsequent transcription and construction of the discussion layer of the dataset. The recording and transcript-verification procedures are described in the following section.

### 2.12 Transcription and verification

The online research tumour-board sessions were recorded and automatically transcribed using the automatic speech-recognition functionality of SaluteJazz (version 26.61.10). The automatically generated Russian-language transcripts were treated as intermediate machine-generated records and were not considered sufficiently reliable for direct use as clinical research data.

Two medical residents independently had access to the original meeting recordings and jointly reviewed the automatically generated transcripts. Verification focused on clinically meaningful transcription errors, including medical terminology, drug names, doses and units, anatomical terms, disease stages, clinically significant negations, speaker attribution, and segmentation of utterances.

When a transcript fragment was unclear, inconsistent with the surrounding clinical reasoning, or interpreted differently by the two reviewers, both residents re-examined the corresponding segment of the original recording and discussed the discrepancy until a consensus transcription was reached. Particular attention was given to errors that could materially alter the clinical meaning of an utterance.

This verification step was necessary because the general-purpose speech-recognition system was not specifically optimised for specialised medical terminology and occasionally produced semantically incorrect renderings of medical terms. The medically reviewed transcript therefore served as the authoritative textual representation of the research tumour-board discussion.

The verified transcript retained speaker attribution and the temporal order of the discussion, allowing preservation of the sequence of arguments, counterarguments, clarifications, and changes in professional position during multidisciplinary interaction.

The raw automatically generated transcripts were retained as intermediate provenance and quality-control records but are not treated as the authoritative discussion layer of Dataset version 1.0. The verified transcripts are currently being used to prepare the English-language dataset representation, as described in the translation procedure below.

### 2.13 Final tumour-board decision

During each research tumour-board discussion, the three specialist annotators presented, defended, and, where appropriate, refined their independently prepared positions. The tumour-board chair moderated the discussion, considered the competing arguments and supporting evidence, and was responsible for formulating the final research decision for each case.

The final decision was formulated during the same tumour-board session after the relevant diagnostic and treatment positions had been discussed. The chair weighed the strengths and limitations of the specialist arguments and determined the interpretation and management strategy considered best supported by the information available within the research workflow.

The three specialist annotators did not subsequently vote on or formally approve the final wording. Their contribution to this stage consisted of presenting their clinical reasoning, responding to counterarguments, and providing the specialty-specific expertise considered by the chair.

Where applicable, the final tumour-board decision included the preferred diagnostic interpretation and disease stage, evidence considered most relevant, the proposed diagnostic and treatment strategy, treatment sequence, conditions for modification of the strategy, response-assessment considerations, and remaining uncertainty.

The final tumour-board decision constituted a separate data layer and was not an edited version of any individual specialist annotation. It represents the final decision formulated by the chair after multidisciplinary discussion and should not be interpreted as an absolute clinical ground truth.

Because the research tumour boards were conducted retrospectively and exclusively for dataset construction, these final decisions did not influence the clinical management or outcomes of the included patients.

### 2.14 Structural assessment of externalised clinical reasoning

To assess whether the specialist records contained explicitly documented clinical reasoning rather than only clinical conclusions, sections B, C, and D were evaluated using a predefined 14-item structural rubric. The assessment was designed to measure the explicitness and structural completeness of externalised reasoning and was not intended to evaluate physician competence, clinical correctness, or adherence to treatment guidelines.

Section B contained four criteria assessing whether the physician explicitly formulated clinical problems or syndromes, linked them to patient-specific supporting evidence, described relevant causal or mechanistic relationships, and considered related but unsupported alternatives.

Section C contained six criteria assessing the formulation of competing hypotheses, relative probability, arguments supporting and opposing each hypothesis, links between hypotheses and verification steps, and consideration of clinically important high-risk alternatives.

Section D contained four criteria assessing whether the proposed diagnostic and treatment strategy specified the clinical objective, conditional or scenario-based logic, sufficiently concrete diagnostic or therapeutic actions, and monitoring of effectiveness and safety.

Each criterion was scored on an ordinal scale from 0 to 3 according to the degree to which the corresponding reasoning element was explicitly represented in the text. Section-level scores were subsequently normalised to a 0–10 scale.

An initial structural assessment was generated using ChatGPT Codex 5.5 extra-high (OpenAI). The model was instructed to evaluate only whether the predefined reasoning elements were present and explicitly expressed in the specialist text and not whether the underlying diagnosis, treatment strategy, or clinical conclusion was medically correct.

The model-generated assessments were subsequently reviewed by two independent physicians holding doctoral research degrees. Each reviewer examined the correspondence between the specialist text, the predefined rubric criteria, and the model-generated scores. When the reviewers disagreed with the automated assessment or with one another, the corresponding text and criterion were re-examined and discussed until a consensus assessment was reached. The human-reviewed assessment was treated as the final structural score.

The resulting assessments were used to provide general, non-personalised feedback to the annotators between sequential annotation rounds. Feedback was intended to increase the explicitness, traceability, and structural completeness of subsequent reasoning records rather than to rank the participating physicians or assess their professional competence.

Because feedback was provided between rounds, the annotation process was intentionally iterative. Structural scores across sequential cases should therefore not be interpreted as independent measurements of physician performance, as later records may partly reflect increasing familiarity with the annotation framework.

### 2.15 Ethics approval and participant consent

The study was approved by the Local Ethics Committee of I.M. Sechenov First Moscow State Medical University (Sechenov University), Moscow, Russian Federation (extract from protocol No. 14-26, dated 2 July 2026). The secondary scientific use and de-identification of the source clinical data were performed within the Sechenov University institutional governance framework. All included patients had provided documented consent permitting secondary scientific use of their clinical data.

Dataset version 1.0 will be distributed under controlled access on request under a data use agreement. The detailed access conditions will be reported together with the repository release in an updated version of this preprint and in the journal-submission version.

All participating physicians provided informed consent for the scientific use of their annotations and for recording, transcription, pseudonymised publication, and subsequent research reuse of their contributions to the research tumour-board discussions.

All patient records were de-identified by authorised Sechenov University personnel before being made available to the research annotators. Direct identifiers were removed as described above. The final dataset will undergo an additional disclosure-risk review before deposition.

Incapacitated patients, minors and other vulnerable groups were not included in the study. The co-authors and annotators did not take part in the treatment of the patients whose cases were included in the dataset, and the tumour-board chair did not take part in case selection.

The study was conducted in accordance with the principles of the Declaration of Helsinki.

### 2.16 Dataset freeze and versioning

The dataset described in this manuscript was defined as version 1.0 and restricted to the ten colorectal cancer cases included in the present Data Descriptor. The version-of-record dataset therefore comprises ten patient-level cases, 30 independent specialist reasoning records, ten verified case-level multidisciplinary discussion records, and ten final tumour-board decisions.

Cases collected after completion of the ten-case dataset were not incorporated into version 1.0. Subsequent cases, additional clinical specialties, primary medical images, computer-vision outputs, extensions of the annotation framework, and downstream artificial intelligence experiments are outside the scope of the present release and will be distributed separately where applicable.

Each patient case, source document, selected evidence fragment, specialist reasoning record, tumour-board discussion unit, and final tumour-board decision was assigned a stable research identifier. These identifiers were preserved across all dataset components to maintain linkage and provenance between the different data layers.

Dataset version 1.0 will contain linked original-language and English-language representations of the textual data. The de-identified Russian-language records will constitute the original-language provenance layer, whereas the aligned English-language records will be provided to facilitate international reuse. Corresponding Russian and English records will share stable case-, document-, annotation-, discussion-, and decision-level identifiers.

The version of record will be accompanied by a complete file manifest, data dictionary, machine-readable schema, annotation-protocol version, translation-procedure version, software version, and changelog. File-level checksums will be generated to enable verification of release integrity.

The repository release corresponding to this Data Descriptor will be treated as immutable. Corrections or subsequent extensions affecting the released data will be issued as separately versioned releases rather than silently modifying version 1.0.

The versioning and metadata strategy was designed to support FAIR principles for data reuse [17].

### 2.17 Translation and language harmonisation

The source clinical records, specialist annotations, verified tumour-board discussion transcripts, and final tumour-board decisions were originally generated in Russian.

To facilitate international reuse, the textual components of Dataset version 1.0 are currently being translated into English by a physician-scientist proficient in both Russian and English and familiar with oncological terminology and the clinical context represented in the dataset.

Translation is being performed at the level of individual source documents, source-linked physician annotations, complete specialist reasoning records, verified tumour-board discussion transcripts, and final tumour-board decisions. Stable research identifiers are preserved during translation so that each English-language record can be traced directly to the corresponding de-identified Russian-language record.

The objective of translation is to preserve clinical meaning rather than to reproduce the source text through literal word-for-word correspondence. Particular attention is given to medical terminology, anatomical terms, disease stage, drug names, treatment regimens, temporal relationships, clinically meaningful negation, expressions of uncertainty, competing hypotheses, benefit–harm considerations, and conditional diagnostic or treatment decisions.

The original Russian-language records are retained without replacement by the translated versions and constitute the authoritative provenance layer of the dataset. English-language records will be provided as an aligned derived layer intended to improve accessibility and international reuse.

Any ambiguity identified during translation is resolved by comparison with the corresponding de-identified Russian-language source record and the clinical context of the case. Translation is not used to introduce new clinical interpretations, modify specialist conclusions, or harmonise disagreements between physicians.

The translation procedure will be versioned as part of Dataset version 1.0, and the language of origin, translation status, and corresponding source identifier will be recorded in the dataset metadata.

## 3 Data Records

Dataset version 1.0 is organised as a set of linked data layers representing the progression from source clinical information to independent specialist reasoning, multidisciplinary tumour-board discussion, and the final tumour-board decision.

The version-of-record release is defined to include ten patient-level cases of histologically confirmed colorectal cancer. Each case is linked to three independently generated specialist reasoning records, one medically verified case-level multidisciplinary discussion, and one final tumour-board decision. Stable research identifiers connect the corresponding patient case, source documents, selected evidence fragments, specialist annotations, discussion records, and final decision.

The source clinical records and research annotations were originally generated in Russian. Dataset version

1.0 is designed to contain linked original-language and English-language representations of the textual data. The de-identified Russian-language records constitute the provenance layer, whereas aligned English-language translations are currently being prepared to facilitate international reuse. Corresponding Russian and English records will share stable identifiers and will be treated as two language representations of the same underlying data object rather than as independent observations.

The principal scientific data objects included in dataset version 1.0 are summarised in Table 1. Exact counts of source documents and source-linked evidence fragments will be calculated automatically from the frozen release before repository deposition.

**Table 1:** Principal data records included in dataset version 1.0.

| Data object | Format | Content | Count |
| --- | --- | --- | --- |
| `metadata/cases.csv` | CSV | Case-level metadata for the ten completed colorectal cancer cases. | 10 |
| `metadata/annotators.csv` | CSV | Pseudonymised metadata for the three specialist annotators and the tumour-board chair, including specialty, experience category, institutional affiliation category, and study role. | 4 |
| `metadata/source_documents.csv` | CSV | Manifest of the de-identified source clinical documents linked to individual patient cases. | – |
| `source_records/` | Mixed | Paired de-identified Russian-language source clinical records and their aligned English-language translations. | – |
| `source_spans/` | JSON Lines | Selected evidence fragments linked to physician interpretations. Russian-language source spans and aligned English-language representations retain common provenance identifiers. | – |
| `specialist_reasoning/` | JSON Lines | Thirty independent specialist reasoning records, with linked Russian-language originals and English-language translations. | 30 |
| `verified_discussions/` | JSON Lines | Ten medically reviewed case-level tumour-board discussion transcripts with speaker attribution and temporal ordering, together with aligned English-language translations. | 10 |
| `final_tumour_board_decisions/` | JSON Lines | Ten chair-formulated final research decisions produced after multidisciplinary discussion, with linked Russian-language originals and English-language translations. | 10 |
| `reasoning_structured_scores.csv` | CSV | Fourteen-item structural assessments of sections B, C, and D, including the initial ChatGPT Codex 5.5 extra-high assessment and the final physician-reviewed scores. | – |

The repository release will additionally contain a README file, a data dictionary, a machine-readable schema, translation metadata, the versioned annotation manual, the prompt used for LLM-assisted preparation of discussion questions, release and validation reports, a changelog, a file manifest, and file-level checksums. These supporting materials describe the structure and provenance of the released records but are not independent clinical data objects.

The complete intermediate set of LLM-generated candidate discussion questions was not systematically archived during data collection and is therefore not represented as a separate reusable data layer. The prompt used for question generation is retained as methodological provenance, whereas the questions actually introduced during the tumour-board sessions can be identified within the verified discussion records where they were spoken.

Raw automatically generated Russian-language automatic speech-recognition transcripts were retained as intermediate provenance and quality-control materials. Because these transcripts contain recognition errors involving specialised medical terminology and were not considered authoritative clinical text, the principal discussion layer of dataset version 1.0 consists of the transcripts that underwent review by two medical residents. The raw ASR outputs are not treated as an additional independent clinical data layer.

### 3.1 Identifiers and linkage

The dataset uses stable research identifiers to connect data objects across the different stages of the data-generation workflow. The identifier architecture was designed to preserve provenance from the patient-level case through source evidence, independent specialist reasoning, multidisciplinary discussion, and the final tumour-board decision.

Each patient-level case is assigned a unique case_id. The ten cases included in dataset version 1.0 are represented by identifiers corresponding to cases 001–010. The research identifiers are not derived from medical-record numbers or other patient identifiers.

Each source clinical document is assigned a unique document_id linked to the corresponding case_id. Individual evidence fragments selected from a source document are assigned a source_span_id. A source span retains a reference to both its parent document_id and case_id, allowing a physician interpretation to be traced back to the clinical document from which the supporting evidence originated.

Each participating physician is represented by a stable pseudonymous annotator_id. The three independent annotators correspond to medical oncology, surgical oncology, and radiation oncology, while the tumour-board chair is represented by a separate identifier and is not classified as an independent annotator.

Each complete specialist annotation of one patient case is assigned a reasoning_record_id. The record is linked to the corresponding case_id and annotator_id and contains the structured annotation fields, source-linked interpretations, diagnosis or problem formulation, recommendations, individual final judgement, and physician-authored questions for multidisciplinary discussion.

Individual evidence-linked annotations within a reasoning record retain the corresponding source_span_id where supporting evidence was explicitly attached. This enables distinction between information originating directly from the patient record, external supporting evidence used by the physician, and the physician’s own interpretation or inference.

Each multidisciplinary discussion is assigned a discussion_id linked to the corresponding case_id. Individual speaker-attributed transcript units may additionally receive an utterance_id, preserving the order of statements within the discussion and the pseudonymous identifier or study role of the speaker.

The chair-formulated final research decision for each case is assigned a final_decision_id linked to the corresponding case_id and discussion_id. This relationship makes it possible to trace the final tumour-board decision to the multidisciplinary discussion from which it was derived while preserving the three original specialist records as separate data objects.

Russian-language and English-language representations share the same underlying research identifiers. Language is represented as metadata, for example through a language field with values ru or en, rather than by assigning the translated record a new clinical identity. A source_record_id or equivalent linkage field connects each English-language translation to the corresponding de-identified Russian-language record.

The resulting linkage structure can therefore be represented conceptually as:

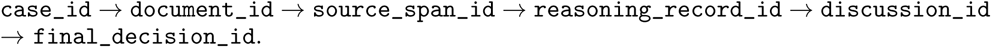

Not every higher-level statement is required to reference a single source span, because sections B–D and the final tumour-board discussion may synthesise information from multiple documents and evidence fragments. The identifier architecture nevertheless preserves all available source links and allows users to distinguish direct source grounding from higher-level clinical synthesis.

## 4 Data Overview

Dataset version 1.0 contains ten completed patient-level cases of histologically confirmed colorectal cancer. The same three specialists—a medical oncologist, surgical oncologist, and radiation oncologist—independently reviewed each case, resulting in 30 specialist–case reasoning records. After completion and locking of the three independent records, each case underwent an online multidisciplinary research tumour-board discussion involving the same three specialists and the same tumour-board chair. The dataset therefore contains ten medically verified case-level discussion records and ten final tumour-board decisions formulated by the chair after multidisciplinary discussion.

The data are hierarchically organised around the patient-level case. Each case may contain multiple deidentified source clinical documents and multiple evidence fragments selected by the participating physicians. These source-level objects are linked to the corresponding specialist reasoning records, which are in turn linked to the multidisciplinary discussion and the final tumour-board decision. The 30 specialist records therefore represent repeated assessments of ten patient cases by the same three physicians and should not be interpreted as 30 independent patients or 30 independent annotators.

The source clinical material, specialist annotations, verified discussion transcripts, and final tumour-board decisions were originally generated in Russian. Dataset version 1.0 retains these de-identified Russian-language records as the provenance layer and will provide aligned English-language translations for international reuse. Russian and English representations of the same underlying data object will share stable research identifiers.

The principal dataset components are summarised in Table 2.

**Table 2:** Overview of the principal data objects in dataset version 1.0.

| Data object | Count | Description |
| --- | --- | --- |
| Patient-level cases | 10 | Completed cases of histologically confirmed colorectal cancer selected for multidisciplinary research review. |
| Independent specialist reasoning records | 30 | Three independently generated records per case, produced by the same medical oncologist, surgical oncologist, and radiation oncologist. |
| Verified multidisciplinary discussion records | 10 | One medically reviewed case-level transcript per patient case, derived from the recorded online research tumour-board discussion. |
| Final tumour-board decisions | 10 | One final research decision per case, formulated by the tumour-board chair after multidisciplinary discussion. |
| Specialist annotators | 3 | The same three specialists independently reviewed all ten patient cases. |
| Tumour-board chair | 1 | The same chair moderated all three research tumour-board sessions and formulated the final tumour-board decisions. |
| Source clinical documents | – | De-identified clinical documents provided by Sechenov University and linked to the corresponding patient cases. |
| Source-linked evidence fragments | – | Evidence fragments selected by physicians and linked to their clinical interpretations across the structured annotation fields. |
| Languages | 2 | De-identified Russian-language provenance records and aligned English-language translations. |

The ten cases were collected over three sequential annotation rounds comprising cases 001–003, 004–006, and 007–010. Because general feedback on the explicitness and structural completeness of documented reasoning was provided between rounds, later specialist records may reflect increasing familiarity with the annotation framework. Dataset version 1.0 should therefore be interpreted as an iteratively generated reasoning dataset rather than as a cross-sectional assessment of physician performance.

## 5 Technical Validation

Technical validation was designed to assess the completeness, traceability, structural consistency, transcription quality, and interpretability of the released data rather than to establish the clinical superiority of any individual specialist opinion or final tumour-board decision. Because dataset version 1.0 contains only ten patient-level cases, several validation procedures were applied to the complete dataset rather than to a sampled subset.

### 5.1 Completeness and relational integrity

Dataset version 1.0 contains ten patient-level cases. For every case, three complete independent specialist reasoning records are available, corresponding to medical oncology, surgical oncology, and radiation oncology. Each case additionally contains one medically verified case-level multidisciplinary discussion and one final tumour-board decision.

All structured annotation fields used in the study were mandatory. The MedMark AIRI platform did not allow an annotator to complete the corresponding stage while required fields remained unaddressed. When information was unavailable, not applicable, or outside the specialist’s role, the physician was required to record an explicit reason rather than leave the field blank.

The resulting frozen dataset therefore contains 30 complete specialist–case reasoning records, ten verified discussion records, and ten final tumour-board decisions.

For the repository release, machine-readable integrity checks will additionally verify uniqueness of primary identifiers, validity of links between cases and associated records, permitted controlled values, UTF-8 encoding, absence of unresolved internal platform identifiers, and consistency between the file manifest and the released data objects. File-level checksums will be generated for the version-of-record release.

### 5.2 Source-link integrity and fidelity

The annotation platform allowed supporting evidence to be linked to any structured annotation field. Patient-derived evidence was linked to identifiable fragments of the corresponding de-identified source clinical document, whereas external supporting materials, where used, were recorded separately from patient-derived evidence.

Technical source-link integrity is defined by the existence of the referenced case, source document, evidence fragment, and associated specialist record. The identifier architecture described above allows these relationships to be checked automatically in the frozen release.

The clinical interpretation associated with a source fragment represents the specialist’s reasoning and is therefore distinct from the source evidence itself. The dataset preserves this distinction to prevent an inferred interpretation from being represented as if it were explicitly stated in the original medical record.

A complete independent semantic audit of all source-linked interpretations has not yet been performed for the present preprint. Before final journal submission, the source-linked layer will undergo case-by-case clinical review to assess whether each referenced fragment supports the associated interpretation and whether direct source information can be distinguished from higher-level inference.

### 5.3 Transcript verification

All three online research tumour-board sessions were automatically transcribed using the automatic speech-recognition functionality of SaluteJazz. The resulting raw Russian-language transcripts contained recognition errors, particularly for specialised medical terminology, and were therefore treated only as intermediate machine-generated records.

Every tumour-board transcript underwent complete medical review by two medical residents who had access to the corresponding original meeting recording. Review included medical terminology, drug names, doses and units, anatomical terminology, disease stage, clinically meaningful negation, speaker attribution, and segmentation of utterances.

When the two reviewers encountered an unclear or disputed fragment, they re-examined the corresponding recording and discussed the fragment until a single agreed transcription was established. The medically reviewed transcript was treated as the authoritative textual representation of the multidisciplinary discussion.

The verified transcripts preserve speaker attribution and temporal ordering, allowing reconstruction of the sequence of arguments, counterarguments, clarifications, and changes in professional position during the tumour-board discussion.

The raw automatic transcripts were retained as intermediate provenance and quality-control material but are not considered an authoritative clinical data layer. Quantitative comparison of the raw and medically verified transcripts, including the number and type of clinically meaningful corrections, may be reported as an additional validation metric in the final repository release.

### 5.4 Structural assessment of externalised reasoning

The explicit structural content of sections B, C, and D was assessed using the predefined 14-item rubric described in the Methods.

For each of the 30 specialist reasoning records, four criteria were evaluated in section B, six criteria in section C, and four criteria in section D. Dataset version 1.0 therefore contains 420 item-level structural assessments.

Each criterion was scored from 0 to 3 according to the degree to which the corresponding reasoning element was explicitly represented in the specialist text. The resulting assessment measured the visibility and structural completeness of documented reasoning and did not evaluate physician competence, diagnostic accuracy, treatment correctness, or adherence to clinical guidelines.

ChatGPT Codex 5.5 extra-high (OpenAI) generated the initial criterion-level assessments. All model-generated assessments were subsequently reviewed by two physicians holding doctoral research degrees. The reviewers examined the specialist text and corresponding rubric criteria and discussed disagreements until a single human-reviewed assessment was reached.

The final human-reviewed scores were used for dataset quality control and for general, non-personalised feedback between annotation rounds. Because the workflow was iterative, changes in structural scores across sequential cases may reflect increasing familiarity with the annotation framework and should not be interpreted as longitudinal changes in physician competence.

Agreement between the initial model-generated scores and the final human-reviewed scores can be quantified in the final release using exact agreement, weighted agreement statistics, mean absolute score difference, and the proportion of model-generated scores modified during human review.

### 5.5 Clinical-content scope and independent review

The dataset was designed to preserve the structure and provenance of clinical reasoning rather than to establish an external gold-standard answer for each patient case. The final tumour-board decision represents the decision formulated by the chair after multidisciplinary discussion and is not presented as absolute clinical ground truth.

The participating specialists had more than ten years of clinical experience in their respective disciplines, and disagreements were explicitly discussed during the research tumour boards. Nevertheless, multidisciplinary discussion alone does not constitute independent validation of clinical correctness.

Before final journal submission, all ten cases will undergo an additional independent review by an oncologist who did not participate in the original annotation or research tumour-board process. The review will examine the fidelity of patient facts to the source documentation, consistency of diagnosis and staging with the available data, the presence of unsupported patient-specific statements, clinically significant treatment or dosing errors, and consistency between the documented discussion and final tumour-board decision.

The purpose of this independent review will be to identify potential clinical-content errors and limitations rather than to replace the existing specialist reasoning or create a new definitive ground truth.

### 5.6 De-identification and disclosure-risk validation

Initial de-identification was performed by authorised Sechenov University personnel within the institution’s protected information environment before the material was uploaded to the research annotation platform. Direct personal identifiers were removed as described in the Methods, and the prepared clinical material underwent an additional review before annotation.

Before public deposition of dataset version 1.0, the complete release package will undergo a final disclosure-risk audit by authorised Sechenov University personnel. This audit will cover the de-identified clinical text, file metadata, internal technical identifiers, healthcare-organisation references, contact information, document identifiers, and combinations of indirect identifiers capable of increasing the risk of re-identification.

Original clinical-event dates are retained because temporal relationships are relevant to retrospective clinical reasoning. Their retention will therefore be explicitly considered as part of the final disclosure-risk assessment.

The completeness of identifier removal was repeatedly checked by several physicians. Together with the small size of the dataset (ten cases), this leads the authors to assess the risk of re-identification as very low; controlled access and the data use agreement provide additional protection.

Only the dataset version approved for scientific data sharing under controlled access following this final audit will be deposited as the version of record.

### 5.7 Translation fidelity

The Russian-language records constitute the original provenance layer of Dataset version 1.0. Aligned English-language translations are currently being prepared to facilitate international reuse and are not treated as independent clinical observations.

Translation from Russian to English is being performed by a physician-scientist proficient in both languages and familiar with the clinical context and oncological terminology represented in the dataset.

The complete English-language layer will subsequently undergo secondary review by an independent bilingual oncologist against the corresponding de-identified Russian-language records. Review will focus on preservation of medical terminology, anatomical descriptions, disease stage, drug names and regimens, temporal relationships, clinically meaningful negation, uncertainty, competing hypotheses, conditional decisions, and the meaning of the final tumour-board decisions.

Discrepancies affecting clinical meaning will be resolved by comparison with the original Russian-language record. Translation review will not be used to harmonise differences between specialists or alter the original clinical reasoning.

Stable research identifiers will link each English-language object to its corresponding Russian-language provenance record, allowing users to verify translation fidelity where required.

## 6 Usage Notes

Dataset version 1.0 contains ten purposively selected patient-level cases of histologically confirmed colorectal cancer and is not intended to represent the full population of patients with colorectal cancer. The dataset should therefore not be used to estimate disease prevalence, frequencies of treatment strategies, or clinical outcomes.

The 30 specialist reasoning records correspond to repeated assessments of the same ten patient cases by the same three physicians. They must not be interpreted as observations from 30 independent patients or 30 independent clinicians. Analyses comparing specialties or individual records should account for the repeated-measures structure of the dataset.

The specialist records represent explicitly externalised clinical reasoning. Absence of a hypothesis, argument, uncertainty, or decision rule from the written annotation does not establish that the physician did not consider it internally. The dataset captures only the portion of reasoning that was explicitly documented or articulated during the research workflow.

The annotation process was iterative. General feedback regarding the explicitness and structural completeness of documented reasoning was provided between annotation rounds. Consequently, later records may reflect increasing familiarity with the annotation framework. Changes across sequential cases should not be interpreted as evidence of improvement in physician competence or clinical performance.

The B–C–D structural scores assess the explicitness and organisation of documented reasoning rather than medical correctness. These scores should not be used to rank physicians, assess professional competence, or determine adherence to clinical guidelines.

The final tumour-board decision for each case was formulated by the chair after multidisciplinary discussion. It was not subsequently approved by formal vote of the three specialist annotators and should not be interpreted as an absolute clinical ground truth. Rather, it represents the final research decision produced within the defined tumour-board workflow.

The source clinical records, specialist annotations, verified discussion transcripts, and final tumour-board decisions were originally generated in Russian. The de-identified Russian-language records constitute the provenance layer of Dataset version 1.0. Aligned English-language translations are currently being prepared to facilitate international reuse. Where interpretation depends on linguistic nuance or translation fidelity, users should refer to the corresponding Russian-language provenance record.

Raw automatic speech-recognition transcripts were not considered authoritative clinical text because the general-purpose transcription system produced errors involving specialised medical terminology. The medically reviewed transcripts constitute the authoritative discussion layer of the dataset.

Primary DICOM examinations and computer-vision outputs were not included in dataset version 1.0. Imaging information is represented only through textual clinical documentation, including radiology reports. Users should therefore not interpret the dataset as a multimodal imaging dataset.

Patient preferences, patient-reported treatment priorities, and patient-defined benefit–harm trade-offs were not systematically captured as a separate structured data layer. The dataset represents multidisciplinary professional decision-making at the patient-case level but should not be described as a complete representation of patient-centred shared decision-making.

The complete intermediate set of LLM-generated candidate discussion questions was not systematically archived. The preserved prompt documents the intended role of the model, while questions that were actually introduced during the tumour-board discussion can be identified from the verified discussion transcript. The dataset should therefore not be used to reconstruct a complete model-question-generation trajectory for every case.

The dataset is not intended to train a general-purpose clinical decision model in isolation. Appropriate reuse scenarios include development and evaluation of methods for source-grounded clinical reasoning, extraction of hypotheses and counterarguments, analysis of cross-specialty differences, identification of unsupported statements, modelling of multidisciplinary discussion, analysis of changes in professional position, evaluation of reasoning-oriented language models, and pilot development of multi-agent clinical decision-support systems.

Any use of the dataset for clinical decision support, model development, or automated medical recommendation requires independent validation in external patient populations and should not be interpreted as evidence of clinical safety or effectiveness.

## 7 Data Availability

At the time of posting this preprint, Dataset version 1.0 is undergoing final de-identification review, English-language harmonisation, technical validation, and repository preparation and is not yet publicly released.

The fixed dataset scope comprises ten patient-level colorectal cancer cases, 30 independent specialist reasoning records, ten medically verified multidisciplinary tumour-board discussion records, ten final tumour-board decisions, and associated provenance and structural-assessment metadata.

Following completion of the final disclosure-risk review, Dataset version 1.0 will be deposited in a persistent research repository with controlled access on request under a data use agreement. The repository URL, data use agreement terms, licence, accession number or DOI, and final file manifest will be added to an updated version of this preprint when the dataset is released.

## 8 Code Availability

MedMark AIRI is proprietary research software developed by the Artificial Intelligence Research Institute (AIRI) and is not distributed as open-source software. The functionality of the platform relevant to this study, including role-based access, case allocation, structured annotation, source-linking, record locking, and transition to multidisciplinary discussion, is documented in the Methods and accompanying figures.

Requests for academic access to additional technical materials or to the frozen research version of MedMark AIRI may be directed to and will be considered in accordance with AIRI research-use and confidentiality policies.

The Dataset version 1.0 repository will provide the machine-readable data schema, data dictionary, export specification, identifier and linkage specifications, prompts, and validation materials required to interpret and validate the released dataset.

The prompt used for LLM-assisted preparation of multidisciplinary discussion questions and the prompt and rubric used for structural assessment of sections B, C, and D will be provided with Dataset version 1.0.

The OpenAI models and SaluteJazz software used during data generation are third-party services and are not redistributed with the dataset.

## 9 Author Contributions

I.V.O. conceived the original concept of representing multispecialty clinical reasoning, disagreement, and tumour-board decision-making as a structured medical dataset and provided overall conceptual leadership for the project.

M.I.S. provided clinical leadership for the project on behalf of Sechenov University, supervised the clinical collaboration, contributed to clinical methodology, and served as chair of all research tumour-board sessions. As tumour-board chair, she moderated the multidisciplinary discussions and formulated the final tumour-board decision for each case.

V.A.G. developed the study design and dataset methodology, coordinated the clinical and technical workflow, supervised dataset construction, contributed to development of the structured clinical-reasoning framework and quality-assessment methodology, coordinated interaction between the clinical and artificial-intelligence components of the project, contributed to medical expert review and interpretation of the dataset architecture, and prepared the original manuscript draft.

D.V.D. contributed to the design and development of the technical research infrastructure and to methodological work related to machine-readable clinical data and artificial-intelligence-assisted processing.

V.D.S. independently reviewed and annotated all ten patient cases from the perspective of medical oncology and participated in the multidisciplinary research tumour-board discussions.

N.N.B. independently reviewed and annotated all ten patient cases from the perspective of surgical oncology and participated in the multidisciplinary research tumour-board discussions.

D.A.B. independently reviewed and annotated all ten patient cases from the perspective of radiation oncology and participated in the multidisciplinary research tumour-board discussions.

A.A.B. contributed to the conceptual and methodological development of the artificial-intelligence-assisted components of the study. He developed and refined the prompting strategy used to generate additional tumour-board discussion questions from the independently completed physician reasoning records, contributed to analysis of model outputs and refinement of the discussion-elicitation workflow, and participated in methodological development and preparation of the manuscript.

N.B.N. contributed to the technical and methodological development of the project at AIRI, including development of the data workflow, machine-readable representation of clinical information, and integration of research components required for dataset construction.

I.R.B. contributed to the development and refinement of the clinical-reasoning methodology and to the design and technical configuration of the workflow used to elicit, structure, and collect clinical reasoning within the MedMark AIRI platform.

I.I.P. contributed to critical review of the manuscript, application of artificial-intelligence methods within the experimental workflow, analysis of AI-assisted components of the study, and preparation and quality control of dataset materials.

A.V.N. contributed to medical expert review, methodological development, and evaluation of the clinical and structural validity of the resulting dataset.

A.V.G. contributed to the design, development, and implementation of the MedMark AIRI annotation platform and the technical workflow used for dataset construction. She contributed to the implementation of role-based access, case allocation, structured annotation, source-linking, record management, and technical support of the clinical annotation workflow.

All authors critically reviewed the manuscript, approved the final version, and accept responsibility for the contributions attributed to them.

## 11 Acknowledgements

The authors gratefully acknowledge all clinicians, researchers, engineers, data specialists, and administrative staff who contributed to the development, implementation, and refinement of this project.

We thank the staff of Sechenov University who supported the identification, preparation, and transfer of the clinical cases and who performed de-identification of the source clinical materials within the protected institutional information environment. We are particularly grateful to the clinical and administrative teams who facilitated access to the source data and supported the organisational procedures required for construction of the dataset.

The authors thank the medical residents who contributed to verification of the automatically generated tumour-board transcripts, including review of specialised medical terminology, speaker attribution, and clinically meaningful transcription errors.

We also thank the physicians and research staff who contributed to quality control of the clinical materials, methodological discussions, and refinement of the structured clinical-reasoning framework.

The authors are grateful to the bilingual physician-scientists contributing to the translation of the de-identified Russian-language clinical materials, specialist reasoning records, tumour-board discussions, and final tumour-board decisions into academic medical English, and to the independent bilingual clinical reviewers contributing to verification of translation fidelity against the original Russian-language records.

We thank the software developers, engineers, and research staff of the Artificial Intelligence Research Institute (AIRI) who contributed to the development and operation of the MedMark AIRI platform, technical implementation of the annotation workflow, data infrastructure, and iterative refinement of the methodology.

Finally, we thank all colleagues who contributed through scientific discussion, methodological review, institutional coordination, technical support, or other forms of assistance during the development of the dataset and preparation of this manuscript.

